# Estimated Average Glucose Integration (eAGi) and A1c Prime (A1c’): A Novel Algorithm for Estimating Glycemic Control Over Time

**DOI:** 10.1101/2025.06.26.25330375

**Authors:** Christopher R. Koltz

## Abstract

Type 2 diabetes affects hundreds of millions worldwide and is associated with significant adverse health outcomes. Optimizing glycemic control mitigates risk over time and has been shown to improve cardiovascular outcomes, reduce microvascular injury as well as decrease overall mortality. Diabetes-specific risk is determined not only by glycemic control but duration of exposure which has been historically difficult to quantify in aggregate. The model proposed in this paper computes two long-term diabetes control scores using a novel algorithm that integrates estimated average glucose as a function of time. The calculated scores are called Estimated Average Glucose Integration (eAGi) and A1c Prime (A1c’). Based on previous studies demonstrating that poor glycemic control over time is associated with worse outcomes, eAGi and A1c’ are anticipated to provide a useful, diabetes-specific hazard appraisal for individuals in both clinical and research settings.

## BACKGROUND

Type 2 diabetes is a pathological condition characterized by impaired glucose metabolism resulting in abnormally high blood glucose levels. Chronically elevated glucose is associated with myriad consequences including atherosclerotic cardiovascular disease (ASCVD), peripheral vascular disease, microvascular complications, stroke and increased mortality among others (1,2). Many of these effects are directly mediated by chronic hyperglycemia while others are driven by insulin resistance and complex metabolic dysregulation (3). The impact of hyperglycemia and insulin resistance is compounded with increasing years of exposure (1,2).

Given the impracticality of frequent glucose measurements, the hemoglobin a1c (A1c) assay was developed to approximate average glucose control over time. A1c is reported as a percent of glycated hemoglobin within erythrocytes and reflects glycemic control over the preceding 60-90 days though there are notable exceptions outside the scope of this review (3,4). A1c is typically used as the criteria for diagnosing diabetes at a threshold of 6.5% and is subsequently used to assess longitudinal glycemic control. The term prediabetes is reserved for A1c values of 5.7% - 6.4% reflecting hyperglycemia but not meeting the criteria for diabetes (4). The relationship between A1c and estimated average glucose (eAG) is defined by the following equation: A1c = (eAG + 46.7)/ 28.7. Conversely, eAG can be converted to A1c using the formula: eAG = (28.7 x A1c) – 46.7. Estimated average glucose is reported in mg/dL and is derived from A1c (4). Only A1c is a direct measurement.

The model proposed in this paper quantifies long-term glycemic control by calculating the area under the curve (AUC) of eAG over time from sequential A1c measurements. The AUC of the eAG-time plot is referred to as eAGi and the derived, weighted average is referred to as A1c prime (A1c’). eAGi is calculated using the trapezoidal rule of integration with each subinterval (in months) graphically represented as a trapezoid, triangle, rectangle or straight line. The subinterval areas are summed, yielding an AUC from time zero to a final time point (Figure 1, Equation 1). The trapezoidal method of integration is particularly well suited for this application given that the slope of y = *f*(x) is constant within any particular subinterval within the eAGi model (Figure 2).

**Figure 1.**
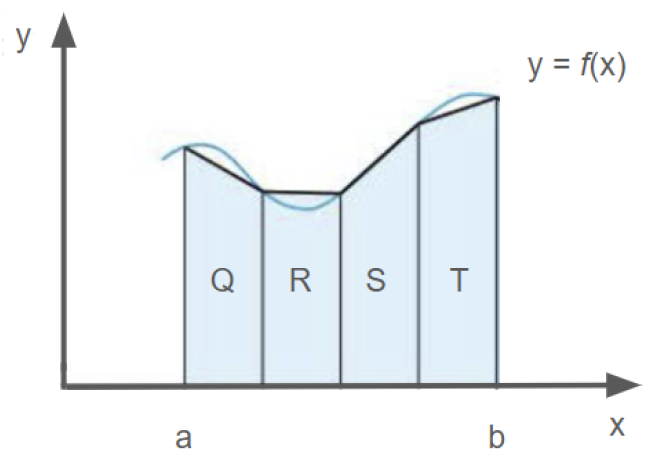
Graphical representation of the trapezoidal rule for integration. For the function y = *f*(x), the integral between a and b is represented by trapezoids labeled Q,R,S,T. Summing the areas of the individual trapezoids yields an approximation of the AUC between a and b.

**Figure 2.**
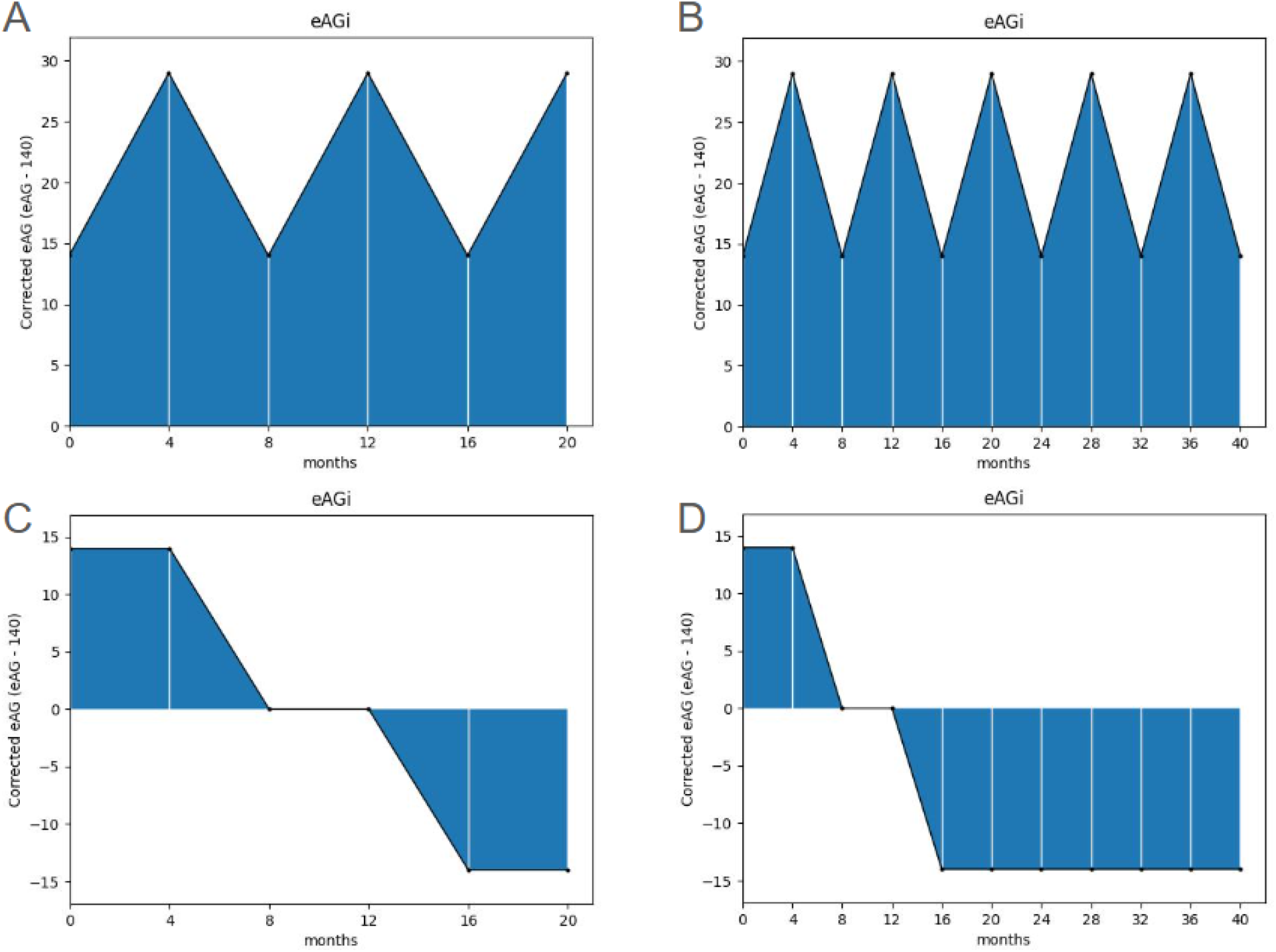
eAGi integrals using test data from Table 1. eAGi is shown in shaded areas. x-axis in months, y-axis in corrected eAG (A) alternating A1c values of 7.0% and 7.5% over 20 months. (B) Same pattern as panel A extended to 40 months. (C) Initial A1c of 7.0% followed by improvement to an A1c of 6.5% with eventual A1c nadir of 6.0% and remaining stable. The total duration is 20 months. (D) Same pattern as panel C initially with a persistent A1c nadir of 6.0% extended to 40 months.

## RESEARCH DESIGN AND METHODS

Four eAGi simulations (Figure 2) were run using fabricated test data (Table 1) to demonstrate the effect of eAG and time on eAGi and A1c’. The derivation for eAGi is shown below (Equation 1).

**Table 1.** Test data for simulations A-D.

|  |  |  |  |  |  |  |  |  |  |  |  |  |
| --- | --- | --- | --- | --- | --- | --- | --- | --- | --- | --- | --- | --- |
| Simulation A | month | 0 | 4 | 8 | 12 | 16 | 20 | .. | .. | .. | .. | .. |
|  | duration* | 0 | 4 | 4 | 4 | 4 | 4 | .. | .. | .. | .. | .. |
|  | A1c | 7 | 7.5 | 7 | 7.5 | 7 | 7.5 | .. | .. | .. | .. | .. |
|  | eAG | 154 | 169 | 154 | 169 | 154 | 169 | .. | .. | .. | .. | .. |
|  | corrected eAG† | 14 | 29 | 14 | 29 | 14 | 29 | .. | .. | .. | .. | .. |
| Simulation B | month | 0 | 4 | 8 | 12 | 16 | 20 | 24 | 28 | 32 | 36 | 40 |
|  | duration* | 0 | 4 | 4 | 4 | 4 | 4 | 4 | 4 | 4 | 4 | 4 |
|  | A1c | 7 | 7.5 | 7 | 7.5 | 7 | 7.5 | 7 | 7.5 | 7 | 7.5 | 7 |
|  | eAG | 154 | 169 | 154 | 169 | 154 | 169 | 154 | 169 | 154 | 169 | 154 |
|  | corrected eAG† | 14 | 29 | 14 | 29 | 14 | 29 | 14 | 29 | 14 | 29 | 14 |
| Simulation C | month | 0 | 4 | 8 | 12 | 16 | 20 | .. | .. | .. | .. | .. |
|  | duration* | 0 | 4 | 4 | 4 | 4 | 4 | .. | .. | .. | .. | .. |
|  | A1c | 7 | 7 | 6.5 | 6.5 | 6 | 6 | .. | .. | .. | .. | .. |
|  | eAG | 154 | 154 | 140 | 140 | 126 | 126 | .. | .. | .. | .. | .. |
|  | corrected eAG† | 14 | 14 | 0 | 0 | -14 | -14 | .. | .. | .. | .. | .. |
| Simulation D | month | 0 | 4 | 8 | 12 | 16 | 20 | 24 | 28 | 32 | 36 | 40 |
|  | duration* | 0 | 4 | 4 | 4 | 4 | 4 | 4 | 4 | 4 | 4 | 4 |
|  | A1c | 7 | 7 | 6.5 | 6.5 | 6 | 6 | 6 | 6 | 6 | 6 | 6 |
|  | eAG | 154 | 154 | 140 | 140 | 126 | 126 | 126 | 126 | 126 | 126 | 126 |
|  | corrected eAG† | 14 | 14 | 0 | 0 | -14 | -14 | -14 | -14 | -14 | -14 | -14 |

\* x values
† y values

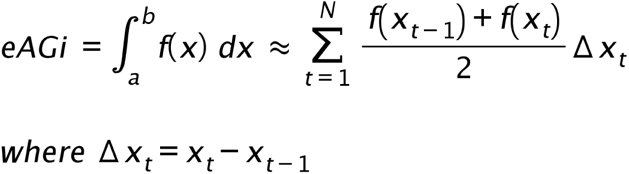

**Equation 1 - Integration by the trapezoidal rule within the eAGi model.**

eAGi is approximately equal to the area summation of all trapezoids (subintervals) between a and b. a = time zero, b = final time point. N = number of subintervals, t = subinterval(1,2..N). Δx_t_ = subinterval duration in months. *f*(*x*_t_) = corrected eAG.

A1c values were first converted to eAG and then to a corrected eAG (eAG - 140) before integration. Corrected eAG values were used to establish an A1c of 6.5% as the reference baseline (y = 0). The final input data includes only paired coordinates for corrected eAG (y value) and subinterval duration (x value). All subintervals in this demonstration are four months long for simplification. The eAGi calculations were coded and executed using Python. The resulting eAGi integrals were then verified against manually performed calculations.

Finally, eAGi results were transformed to their corresponding A1c’ values. The conversion formula is: A1c’ = [(eAGi/total duration) + 140 + 46.7] / 28.7. The value 140 is used to correct the initial eAG manipulation previously described and 46.7 and 28.7 are derived from the A1c to eAg conversion equation: A1c = (eAG + 46.7)/ 28.7. To simplify, eAG values are represented as rounded integers.

## RESULTS

Four hypothetical, proof of concept simulations were run to demonstrate the effect of eAG and time on eAGi and A1c’. Input data is shown in Table 1 and the results for eAGi are graphically displayed in Figure 2. The shaded areas represent eAGi. Subintervals with A1c values of 6.5% are arealess and are depicted as a horizontal line at y = zero (Figure 2, Panels C, D) and serve as a baseline reference for eAGi measurements. All simulations used equal four month subintervals.

Simulation A (Figure 2, Panel A): eAGi = 430, A1c’ = 7.2%. Oscillating pattern between an A1c of 7.0% and 7.5% over 20 months. The eAGi and A1c’ are greater than zero indicating glycemic control is higher than 6.5% on average.

Simulation B (Figure 2, Panel B): eAGi = 860, A1c’ = 7.2%. Same input data and oscillating pattern as simulation A with an extended duration of 40 months. The resulting eAGi is twice that of simulation A however, A1c’ remains the same at 7.2%. Simulation B demonstrates the unique, aggregate duration effect on eAGi but not A1c’.

Simulation C (Figure 2, Panel C): eAGi = zero, A1c ‘= 6.5%. Initial hyperglycemia with an A1c of 7.0% followed by improvement to 6.5% and eventual glycemic nadir of 6.0%. The arealess region in the center of the plot represents a subinterval with an A1c of 6.5% which does not contribute to eAGi. Simulation C demonstrates the additive and regressive nature of eAGi as the early hyperglycemic subintervals are offset by subsequent subintervals with A1c values less than 6.5%.

Simulation D (Figure 2, Panel D): eAGi = - 280, A1c’ = 6.3%. Simulation C is reproduced for the first 20 months with the final A1c nadir of 6.0% extended to 40 months. eAGi becomes negative with a longer duration of glycemic control below an A1c of 6.5%. If this pattern were extended indefinitely, eAGi would progressively become more negative though A1c’ would remain positive and approach a nadir of 6.0% in an asymptotic manner (not shown).

## DISCUSSION

### eAGi as a theoretical predictor of diabetes-related complications

eAGi quantifies long-term diabetes control by integrating sequential glycemic measurements as a function of time. Higher A1c values over time will result in a larger, more positive eAGi as will longer duration of exposure. Ideally, eAGi should be reported in conjunction with the corresponding duration to provide a more precise interpretation. Any time interval can be used for eAGi as long as expressed in months. Fractional intervals such as 1.5 months (∼ 6 weeks) will also produce valid results. Moreover, the zero threshold can be adjusted up or down depending on the desired risk assessment for bespoke objectives.

There is no upper or lower bound for eAGi. It is constrained only by the limits of glycemia compatible with life and the total duration of exposure. In the current model, eAGi will remain zero if A1c values are consistently 6.5% no matter the duration of exposure. A1c values below 6.5% will result in negative numbers for a given subinterval and decrease the composite eAGi. Conversely, A1c values greater than 6.5% will be additive..

The eAGi has potential clinical and research applications. eAGi may help clinicians risk stratify patients and inform management decisions such as escalating and de-escalating therapies as well as aid in perioperative risk assessment. eAGi could also classify research subjects into subgroups for study allocation and analysis. Typically, study participants are grouped together as a single diabetes cohort even though there may be very different diabetes-specific risk profiles based on duration of exposure and long-term glycemic control.

### A1c’ as a complementary metric

A1c’ is intended to be a user-friendly number expressed in a way that is understood by clinicians and patients alike. The resulting value is essentially an average of averages over time (see Methods section for formula). It is worth noting that A1c’ is not interchangeable with eAGi. Unlike eAGi, A1c’ does not directly account for exposure duration resulting in more limited utility. Examples of discordant eAGi and A1c’ are shown in Figure 2 (panel A vs. B). Moreover, A1c’ is bounded by upper and lower limits. To illustrate, A1c’ is always positive unlike eAGi, with a confined range within the limits of the A1c assay. Still, the concept of A1c’ will be familiar to patients and clinicians and provide a meaningful metric to evaluate glycemic control over time.

### Shortcomings of the eAGi model

While there is utility in the proposed eAGi model, there are several limitations to consider. Missing data may produce less accurate results since glycemic variability will not be sufficiently captured. Long subintervals between a1c measurements can result in both overestimation and underestimation depending on the a1c trend. Overestimates are more likely in cases of improving glycemic control and underestimates are more likely with worsening glycemic control. The estimates will only be unaffected when glycemic control is stable or in cases where the rate of change between subintervals is constant.

Moreover, an eAGi of zero corresponds to the standardized baseline and is not meant to imply optimal control. The eAGi results should more correctly be interpreted as glycemic control relative to an eAGi baseline of zero. This point becomes more important when considering the potential use of eAGi as a diabetes-specific risk assessment tool in the future given it does not account for relevant co-morbidities and scores of zero likely do not represent the lowest risk category.

Finally, the presented simulations used rounded integers for the A1c to eAG conversion for simplification. For example, the proper transformation of an A1c of 6.5% to eAG is 139.8 mg/dL not 140 mg/dL. For demonstration purposes, the discrepancy is negligible, especially over short durations with the difference in A1c values being 6.5% and 6.507%, respectively. This variance is below the level of discrimination for the A1c assay.

### Future directions

Higher eAGi and A1c’ values are anticipated to correlate with worse health outcomes based on extensive data examining diabetes related complications as a function of glycemic control and duration of exposure (1,2). Validation of the eAGi model with real-world patient datasets are necessary to further demonstrate its usefulness and provide context.

The manual eAGi computation is impractical at scale but easily managed by capable software. Integrating the eAGi algorithm into electronic medical records (EMR) would provide clinicians a simple and dynamic metric to support point-of-care decisions. Similar, automated risk calculators for metabolic associated liver disease (MASLD) and ASCVD already exist within some EMR platforms. Additionally, the algorithm may prove useful in the research environment by allowing investigators to stratify participants with diabetes by eAGi and/or A1c’ rather than as a single diabetic cohort. This could be employed both prospectively and retrospectively.

## CONCLUSIONS

The eAGi model integrates estimated average glucose as a function of time to calculate two longitudinal diabetes control scores. Higher eAGi values are anticipated to correlate with unfavorable clinical outcomes thereby making eAGi a diabetes-specific *risk* score. Validation studies are needed prior to making this claim, however. A1c’ is derived from eAGi and provides a glycemic, time-weighted average but does not capture absolute exposure therefore having more narrow applicability by comparison.

## Data Availability

All data produced in the present study are available upon reasonable request to the authors

